# PVC Variability and Impact on Meeting Expert Consensus Cutoffs of ≥10,000 PVCs/day and ≥15% PVCs

**DOI:** 10.1101/2024.06.10.24308734

**Authors:** Richard S. Amara, Jason Appelbaum, Ameer Abutaleb, Rama Vunnam, Stephen R. Shorofsky, Jeffrey N. Rottman, Ardit Kacorri, Timm M. Dickfeld

## Abstract

**Background:** Frequent PVCs have been associated with a reversible cardiomyopathy. Cutoffs of ≥10,000 PVCs/day and ≥15% PVCs have been suggested by the 2014 EHRA/HRS/APHRS and 2017 AHA/ACC/HRS Expert Consensus guidelines, respectively, for PVC suppression.

**Methods:** 606 patients with 14 day ZIO® monitor datasets with ≥10,000 PVCs on at least one day were identified (2014 Guidelines Cohort). Of these patients, 325 had at least one day of ≥15% PVCs (2017 Guidelines Cohort). Analysis was performed on these cohorts to investigate the impact of PVC variability on meeting guideline thresholds.

**Results:** Within the 2014 Guidelines Cohort, mean daily PVC burden was 12,188±8,300 [range 0-64,188]. Intra-patient daily PVCs were highly variable (median 3.6-fold change between max and min PVC days (Q1/3: 2.22/10.15) with instances of >10,000-fold change observed. 54.3% and 19.5% of patients had days with <5,000 and <1,000 PVCs, respectively. Even patients with days of 0 PVCs (0.5%) were observed. 72h monitoring detected 69% of patients with ≥10,000 PVCs/24h with an additional 2-4% of patients crossing the threshold each additional day. A bimodal distribution of number of days meeting PVC thresholds/corresponding PVC counts was observed, suggesting a previously unidentified pattern of distinct populations – “low frequency/low PVC” (≤3/14 above-threshold days/mean 12,342 PVCs those days) vs “high frequency/high PVC” (14/14 above-threshold days/mean 24,580 PVCs those days). The 2017 Guidelines Cohort demonstrated similar findings.

**Conclusion:** Daily PVC burdens vary greatly. More sensitive detection of guideline-suggested cut-offs requires prolonged monitoring. Novel potential PVC patterns may allow for better identification of candidates for PVC suppression.

## INTRODUCTION

Premature ventricular contractions (PVCs) are ectopic heart beats which may be present in patients both with and without structural heart disease. While often asymptomatic, they can cause symptoms including palpitations, chest discomfort, and dyspnea. In some cases, PVCs can cause a reduction in left ventricular systolic function, a condition known as PVC-induced cardiomyopathy.^1^ While the exact pathophysiology of PVC-induced cardiomyopathy is yet unclear, studies have demonstrated that the degree of cardiomyopathy may be proportional to 24-hr PVC burden.^2–4^ Additionally, multiple studies have demonstrated that PVC suppression (via antiarrhythmics and/or radiofrequency ablation) can result in reversal of cardiomyopathy and improvement in symptoms.^1–3,5–7^

It is generally thought that a certain PVC burden must be met to induce a cardiomyopathy. Though a definitive cutoff likely does not exist, different guideline recommendations suggest when PVC suppression may be beneficial. The 2014 EHRA/HRS/APHRS Expert Consensus document suggests ≥10,000 PVCs per 24 hours, while the 2017 AHA/ACC/HRS Expert Consensus document suggests ≥15% of daily heart beats.^8,9^ Typically, Holter monitoring has been used to gauge PVC burden, and it is still accepted practice to assess said burden with a single 24-hour Holter monitor. However, there are several prior studies which have shown daily PVCs on Holter monitoring to be quite variable, and it is not clear whether a 24-hour monitoring period provides a confident reflection of long-term PVC burden.^10–13^ The purpose of this study is to investigate daily PVC variability through analysis of 14-day rhythm monitors in a large cohort of patients, in order to gain insight into said variability’s potential impact on meeting established thresholds in the consideration of PVC suppression.

## METHODS

To acquire a number of datasets large enough for meaningful analysis, data was obtained from the national iRhythm commercial data warehouse which houses all ZIO® (iRhythm, San Francisco, USA) monitor datasets. A query was placed for a convenient sample of approximately 1,000 consecutive 14 day ZIO® monitor datasets with at least one day of ≥10,000 PVCs over 24 hours – this resulted in a sample of 1,006 consecutive datasets collected between December 2018 and January 2019, with a wear period of ≥7 days. Only datasets containing the complete 14 days of data (606 datasets) were included in the study. Data points available for analysis included total daily heart beats and total daily PVCs. Patient demographics were not identified. While all patients were referred for rhythm monitoring for a clinical indication, the indications were not specified. Two cohorts of patients were generated – a cohort of patients with at least one day of ≥10,000 PVCs over 24 hours (referred to as the “2014 Guidelines” Cohort), and a sub-cohort of patients with at least one day of ≥15% PVC burden (referred to as the “2017 Guidelines” Cohort). Communication with the IRB of the University of Maryland School of Medicine confirmed that no special IRB approval was required given that only anonymous and deidentified data was used. Research was performed at the University of Maryland Medical Center in Baltimore, Maryland.

The following outcomes of interest were measured and analyzed in these groups:

1. Total PVC counts and PVC burden including average mean, lowest, and highest. PVC burden is expressed as a percentage (total PVC number divided by total heart beats).
2. Intra-patient PVC variability, expressed as fold changes between maximum and minimum PVC days as well as fold changes between consecutive monitoring days.
3. Number of days where PVC thresholds were met and unmet.
4. Number of consecutive monitoring days to reach a PVC threshold.
5. “Tachycardic” and “non-tachycardic” patients and days were identified based on an average HR ≥100 bpm.

### Statistical Analysis

Means, medians, standard deviations, quartiles, and interquartile ranges were identified for variables of interest. Means were compared with one-tailed, paired T-tests. With fold-change computation, days of 0 PVCs were converted to 1 PVC to allow for mathematical analysis (to prevent division by 0). In the comparison of patients with ≤3 days vs all 14 days of ≥10,000 PVCs or 15% PVC burden over 24 hours, two-tailed, unequal variance T-test analysis was performed. “Tachycardic” and “non-tachycardic” patients and days were identified and compared through two-tailed, unequal variance T-test analysis. All statistical analyses were carried out using Microsoft Excel and IBM SPSS. All data analysis was performed in a blinded fashion.

## RESULTS

606 patient datasets with at least one day of ≥10,000 PVCs made up the “2014 Guidelines Cohort” of patients. 325 of these 606 patient datasets had at least one day of ≥15% PVCs, making up a second “2017 Guidelines” sub-cohort of patients. Data for the “2014 Guidelines Cohort” are primarily presented in terms of PVC counts while data for the “2017 Guidelines Cohort” are primarily presented in terms of PVC percentage.

### PVC Counts and ranges

#### 2014 Guidelines Cohort

A total of 8,484 days of data from 606 patients were assessed. Daily PVCs ranged from 0 to 64,188 (Figure 1). The per-patient average mean daily PVC count was 12,188 ± 8,300, significantly higher than the average lowest daily count of 6,356 ± 6,861 and significantly lower than the average highest daily count of 19,851 ± 9,534 (p <0.001 for both). Per-patient mean daily PVC counts are shown in Figure 2a, and per-patient peak day PVC counts are shown in Figure 2b.

**Figure 1:**
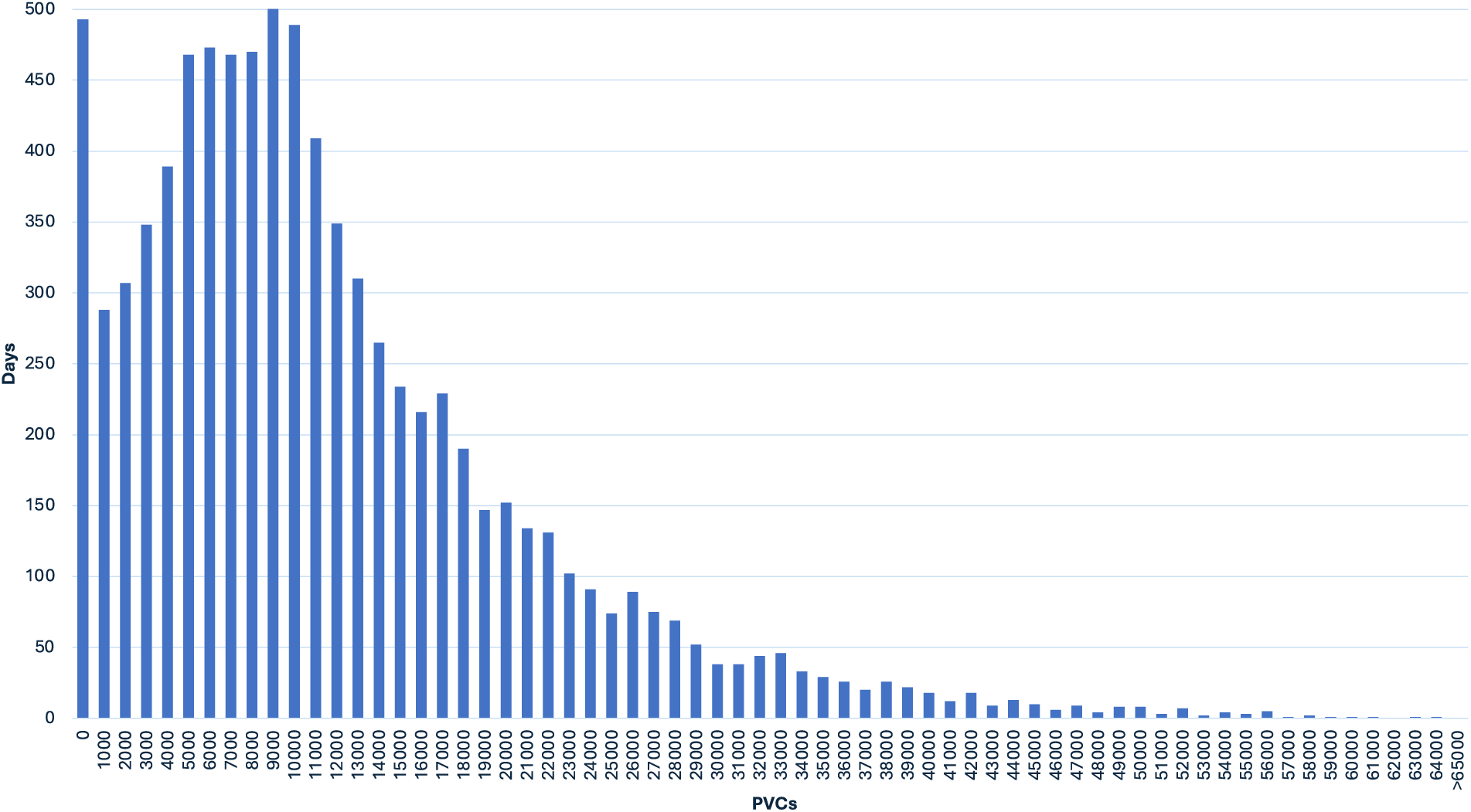
Distribution of PVC Counts (Total Analyzed Days) Histogram depicting the distribution of PVC counts among all analyzed days (8,484 days), with each bar representing a range of 1,000 PVCs.

**Figure 2a:**
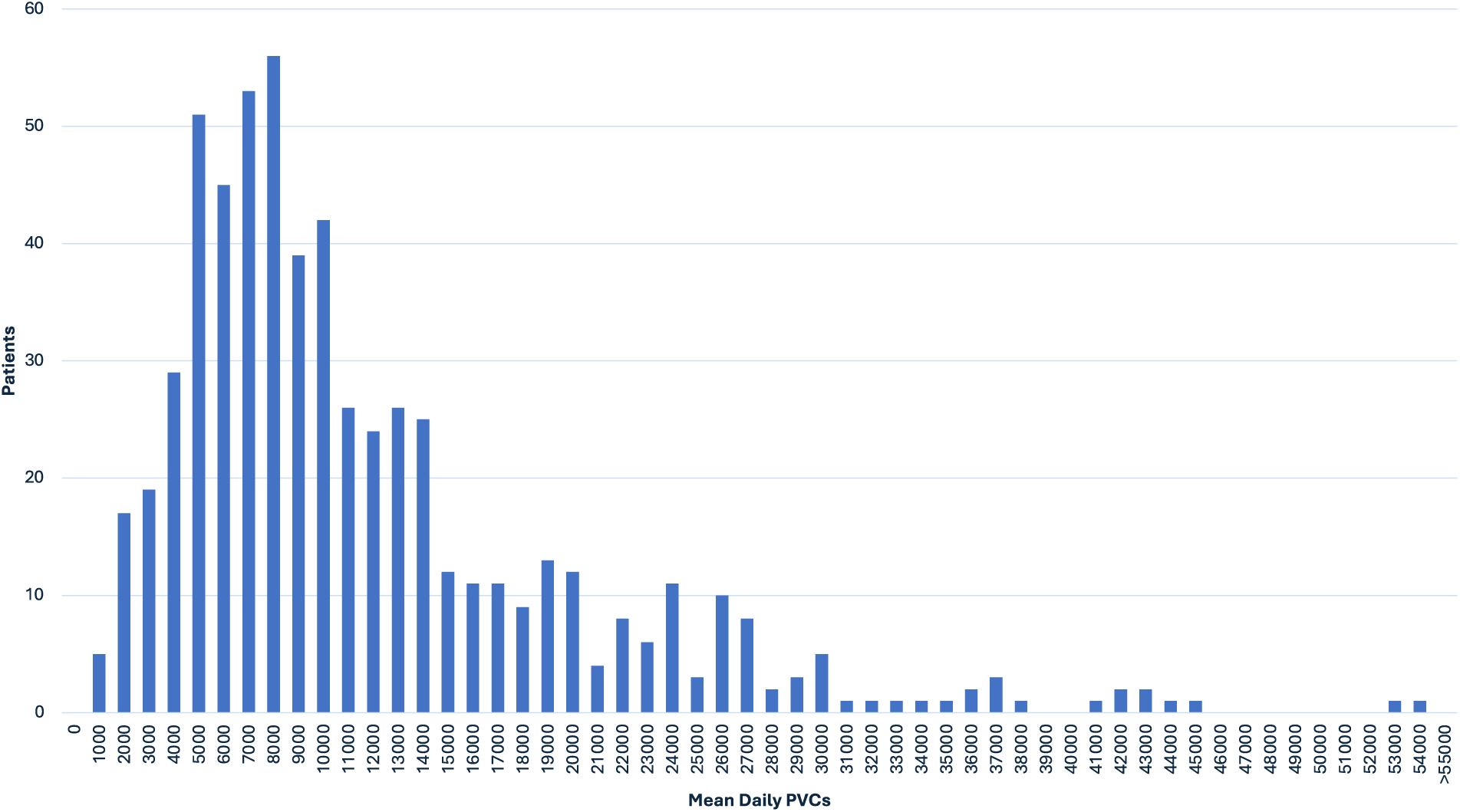
Mean Per-Patient Daily PVC Count (2014 Guidelines Cohort) Histogram depicting the distribution of mean per-patient daily PVC counts (mean PVC count over a 24 hour period, per patient) in the 2014 Guidelines Cohort (≥10,000 PVCs on at least one day), with each bar representing a range of 1,000 PVCs.

**Figure 2b:**
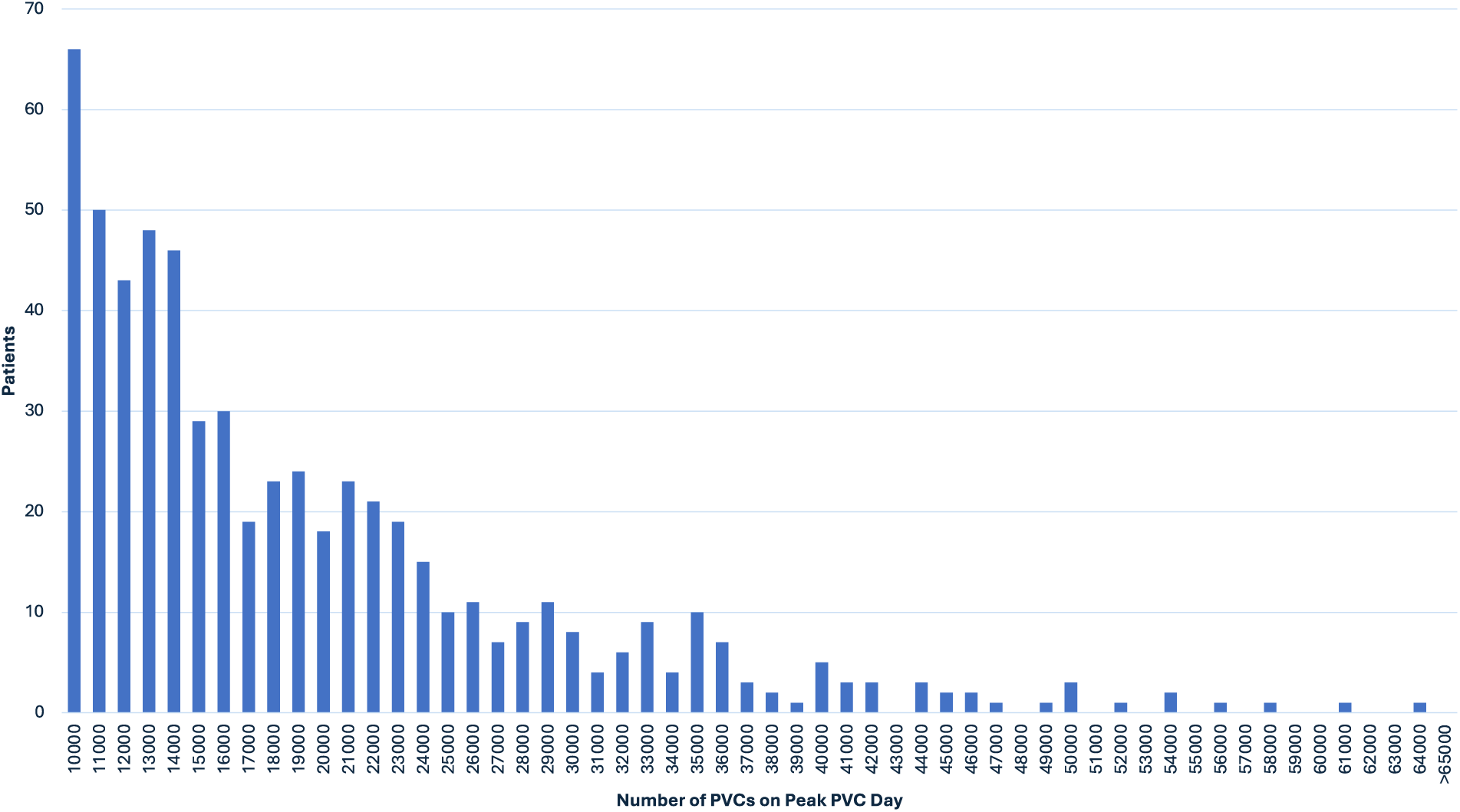
Peak Day of PVCs Per Patient (2014 Guidelines Cohort) Histogram depicting the distribution of the number of PVCs on each patient’s peak day of PVCs in the 2014 Guidelines Cohort (≥10,000 PVCs on at least one day), with each bar representing a range of 1,000 PVCs.

#### 2017 Guidelines Cohort

4,550 days of data from 325 patients were assessed. Daily PVC burden ranged from 0% to 48.1% (Figure 3). The per-patient average mean daily PVC percentage was 15.9 ± 7.8%, significantly higher than the average lowest daily percentage of 9.2 ± 7.7% and significantly lower than the average highest daily percentage of 24.0 ± 7.5% (p <0.001 for both). Per-patient mean PVC percentages are shown in Figure 4a, and per-patient peak daily PVC percentages are shown in Figure 4b.

**Figure 3:**
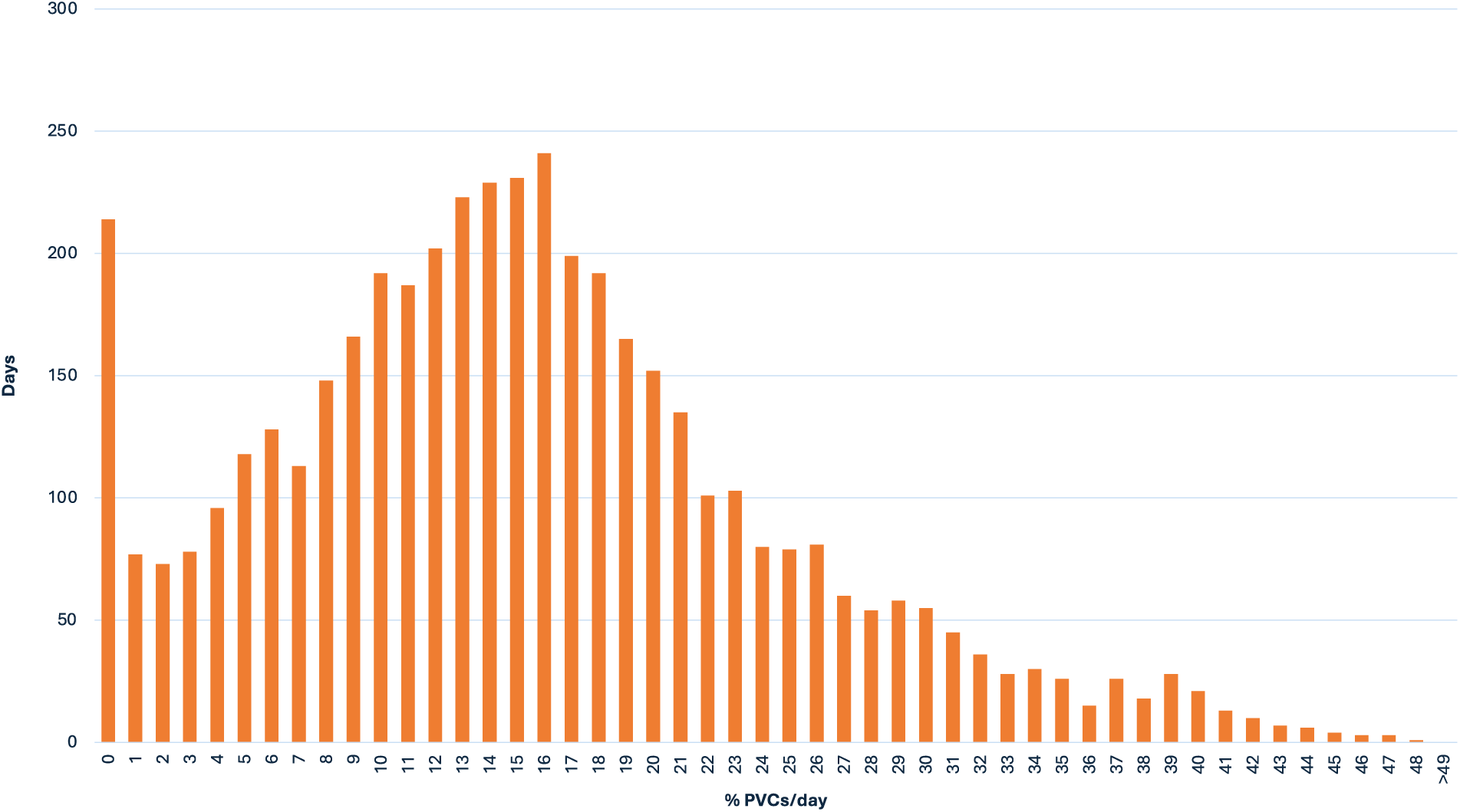
Distribution of PVC Percentage (2017 Guidelines Cohort) Histogram depicting the distribution of PVC counts among days in patients in the 2017 Guidelines Cohort (PVC burden ≥15% on at least one day), with each bar representing a range of 1% PVC burden.

**Figure 4a:**
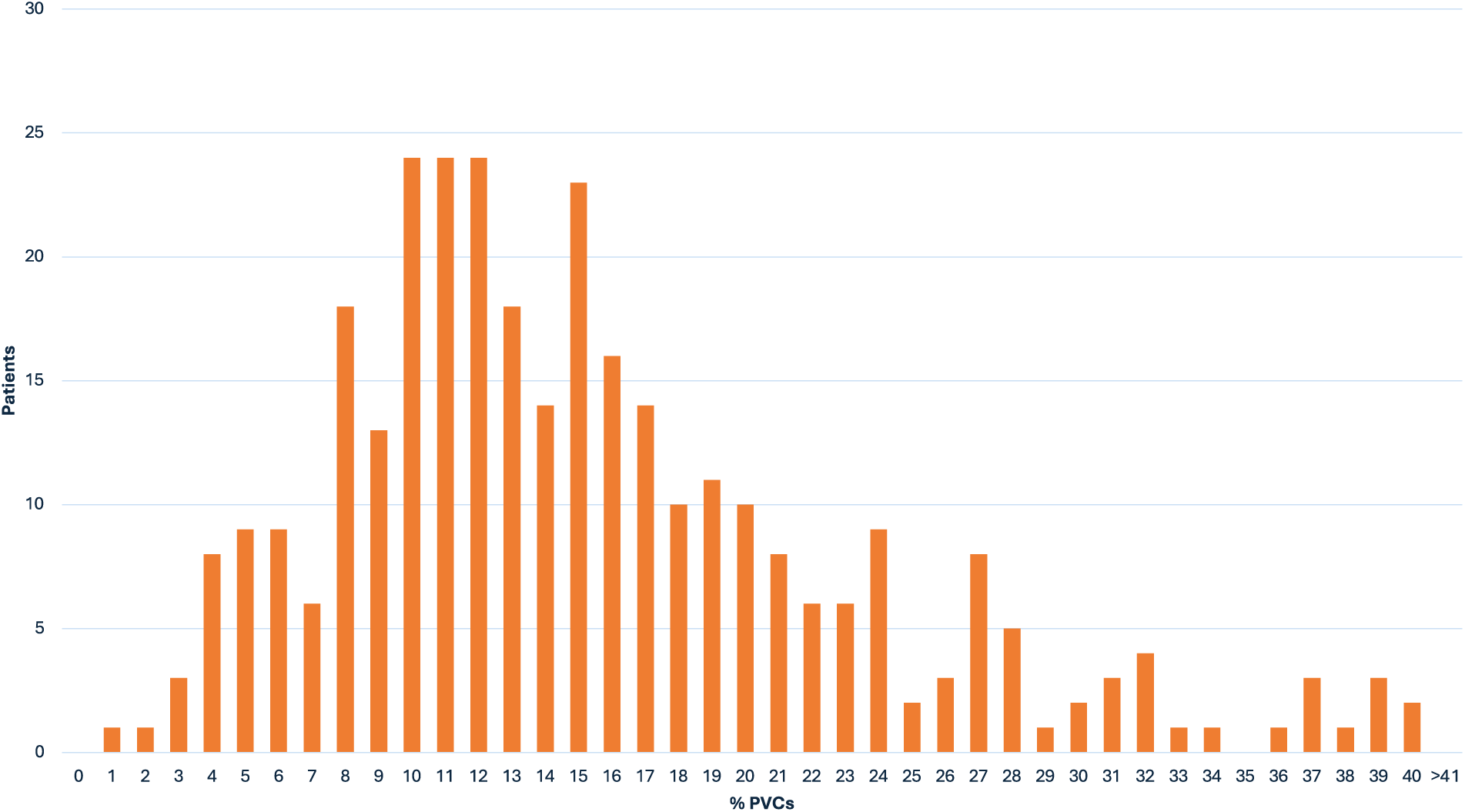
Mean Per-Patient Daily PVC Percentage (2017 Guidelines Cohort) Histogram depicting the distribution of the mean per-patient daily PVC percentage in the 2017 Guidelines Cohort (PVC burden ≥15% on at least one day), with each bar representing a range of 1% PVC burden.

**Figure 4b:**
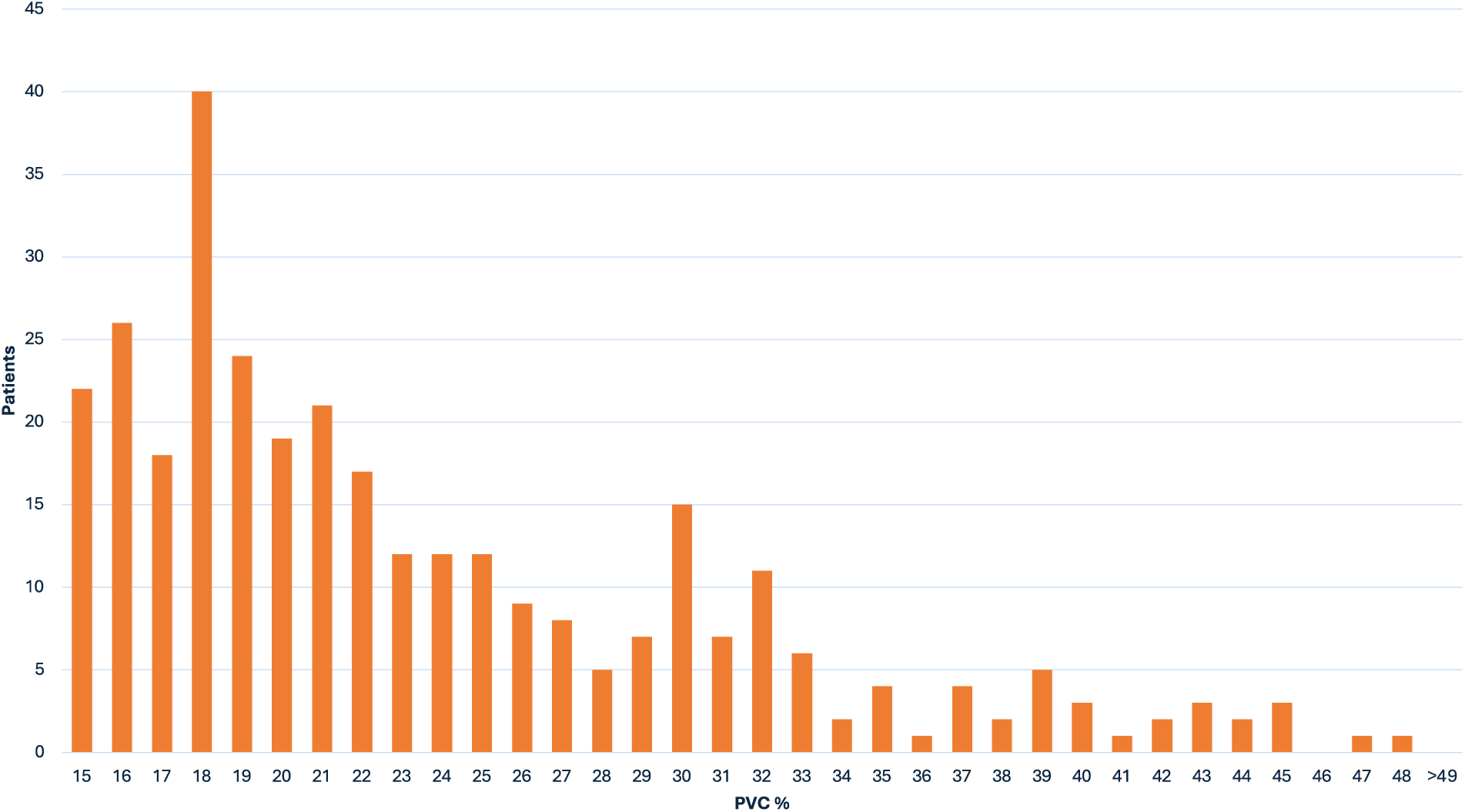
Peak Day PVC Percentage (2017 Guidelines Cohort) Histogram depicting the distribution of the peak day PVC percentage per patient in the 2017 Guidelines Cohort (PVC burden ≥15% on at least one day), with each bar representing a range of 1% PVC burden.

PVC histograms in both cohorts demonstrate that in this PVC-enriched sample population, PVC distribution is not normal. A positive skew is observed whether looking at PVCs from a viewpoint of all days monitored, mean per-patient daily PVCs, or peak per-patient PVC days. When looking at overall days of PVCs, a bimodal distribution is observed. In the 2014 Guidelines Cohort, peaks are observed at 0-1,000 and 5,000-10,000 PVCs (Figure 1). In the 2017 Guidelines Cohort, similar peaks are observed at 0-1% and 10-18% PVCs (Figure 3). This demonstrates that even among patients meeting guideline-suggested PVC cutoffs, there are many days with very few PVCs.

### Intra-patient PVC Variability

#### 2014 Guidelines Cohort

Intra-patient daily PVC counts were highly variable. The intra-patient maximum and minimum day of PVCs varied by a median of 3.60-fold (Q1/3: 2.22/10.15). 111 patients (18.3%) had a fold change of <2, 341 patients (56.3%) had a fold change between 2 and 10, 112 patients (18.5%) had a fold change between 10 and 100, 27 patients (4.5%) had a fold change between 100 and 1,000, 8 patients (1.3%) had a fold change between 1,000 and 10,000, and 7 patients (1.2%) had a fold change of >10,000 (Figure 5).

**Figure 5:**
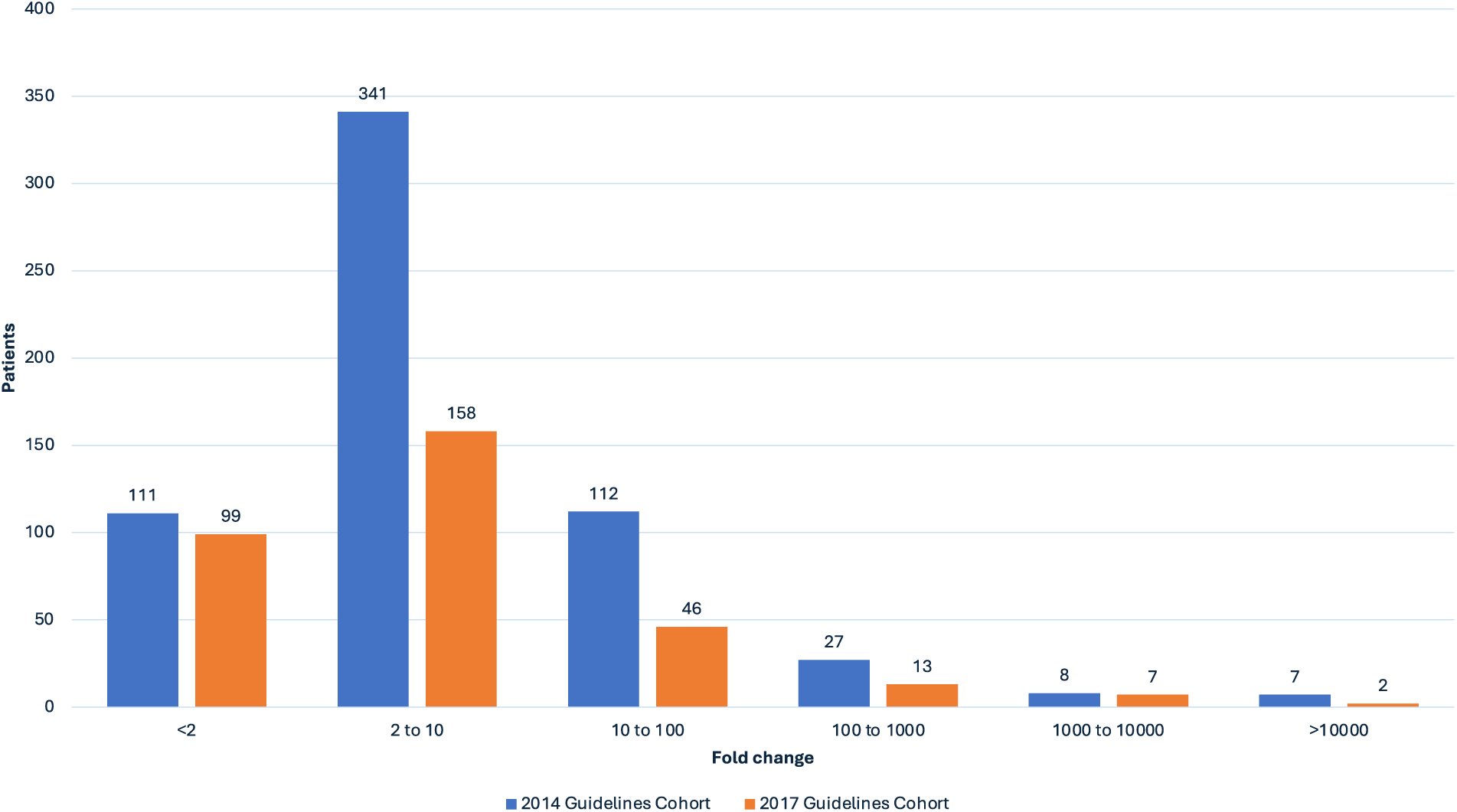
Instances of Fold Change Between Maximum/Minimum PVC Days. Bar chart depicting fold change between maximum and minimum PVC days. Blue bars represent the 2014 Guidelines Cohort (≥10,000 PVCs on at least one day), and orange bars represent the 2017 Guidelines Cohort (PVC burden ≥15% on at least one day). The exact number of patients in each range is noted above each bar.

#### 2017 Guidelines Cohort

Intra-patient daily PVC percentage was similarly highly variable with a relatively similar stratification. The intra-patient maximum and minimum day of PVCs varied by a median of 2.69-fold (Q1/Q3 1.83/6.49). The distribution of maximum to minimum fold change among this cohort is also presented in Figure 5.

### Intra-patient Day to Day PVC variability

#### 2014 Guidelines Cohort

Day to day PVC variability is defined here as PVC changes between subsequent consecutive days. Substantial variability was observed. Day to day PVCs varied by a median of 1.26-fold (Q1/3: 1.10/1.62). Given 14 monitoring days for 606 patients, there were 7,878 day to day instances to assess. Of these 7,878 instances, there were 6,716 (85.3%) instances of <2 fold change, 1,075 (13.7%) instances of between 2 and 10 fold change, 71 (0.90%) instances of between 10 and 100 fold change, 17 (0.22%) instances of between 100 and 1,000 fold change, and 3 (0.04%) instances of a fold change of >1,000. The peak day to day fold change was 4,609 (Figure 6).

**Figure 6:**
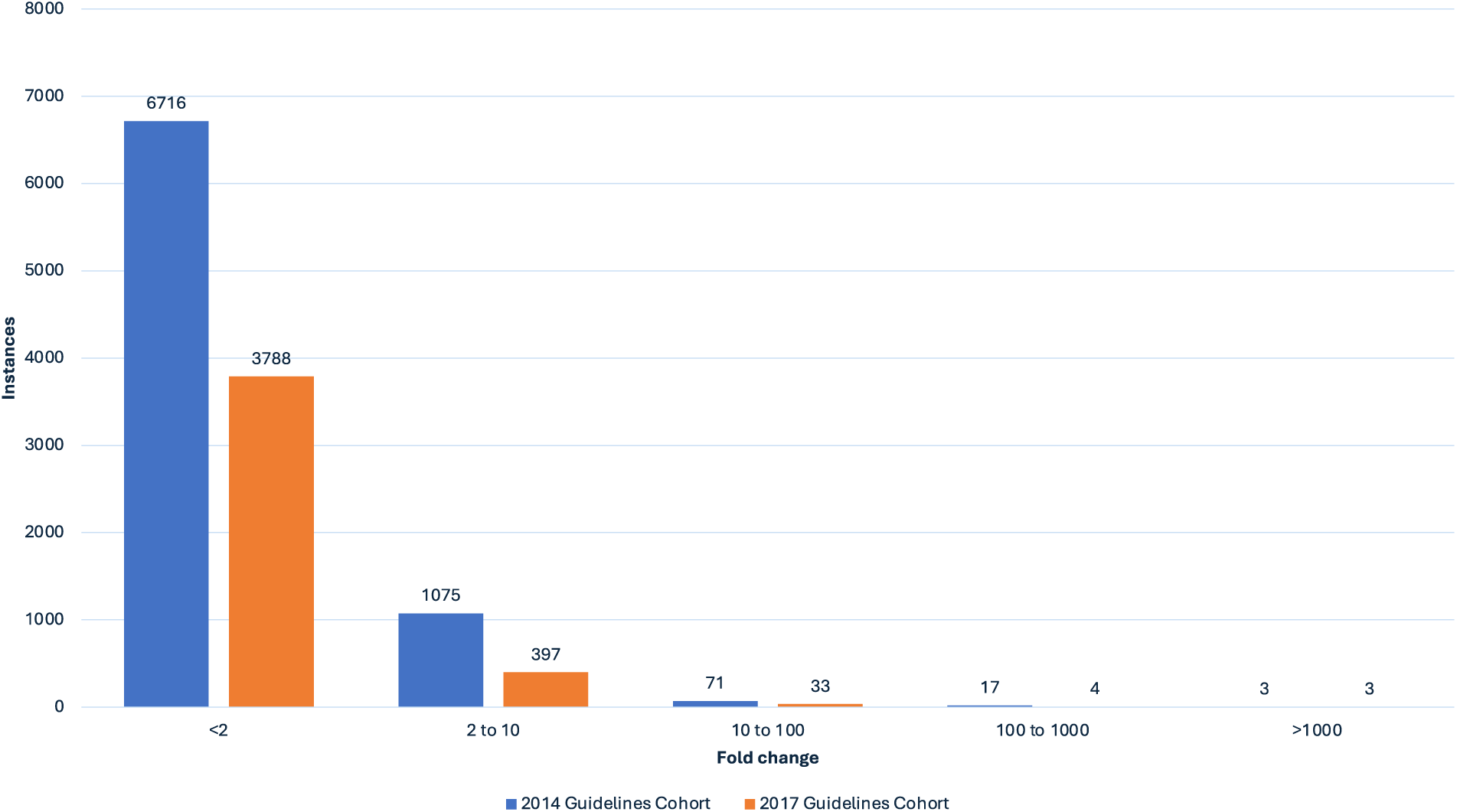
Instances of Day to Day Fold Change. Bar Chart depicting instances of day to day fold change in PVCs. Blue bars represent the 2014 Guidelines Cohort (≥10,000 PVCs on at least one day), and orange bars represent the 2017 Guidelines Cohort (PVC burden ≥15% on at least one day). The exact number of patients in each range is noted above each bar.

#### Day to Day change, 2017 Cohort

Day to day PVCs varied by a median of 1.19-fold (Q1/3: 1.07/1.46). A detailed breakdown of instances of day to day PVC fold changes in this cohort is also provided in Figure 6.

### Days Below Low-PVC Cutoffs

#### 2014 Guidelines Cohort

Significant variability was also evident when assessing how often patients fulfilling guideline criteria had days under the cut-off of 10,000 PVCs. Of the 8,484 days, there were 4 days (0.05%) of 0 PVCs, 52 days (0.61%) of <10 PVCs, 143 days (1.7%) of <100 PVCs, 493 days (5.8%) of <1,000 PVCs, 1,825 days (21.5%) of <5,000 PVCs, and 4,208 days (49.6%) of <10,000 PVCs. Of the 606 patients, this equated to 3 patients (0.5%) with one or more day of 0 PVCs, 12 patients (2.0%) with one or more day of <10 PVCs, 37 patients (6.1%) with one or more day of <100 PVCs, 118 patients (19.5%) with one or more day of <1,000 PVCs, 329 patients (54.3%) with one or more day of <5,000 PVCs, and 482 patients (79.5%) with one or more day of <10,000 PVCs (Table 1).

**Table 1:** Days and Patients under certain PVC counts (2014 Guidelines Cohort). This table depicts how commonly there were days below certain PVC count thresholds, as well as how many patients had at least one day below certain PVC count thresholds.

| **PVCs** | **0** | **<10** | **<100** | **<1000** | **<5000** | **<10000** |
| --- | --- | --- | --- | --- | --- | --- |
| Days (of 8,484) | 4 | 52 | 143 | 493 | 1825 | 4208 |
| Days (%) | 0.05% | 0.61% | 1.69% | 5.81% | 21.51% | 49.60% |
| Patients (of 606) | 3 | 12 | 37 | 118 | 329 | 482 |
| Patients (%) | 0.50% | 1.98% | 6.11% | 19.47% | 54.29% | 79.54% |

#### 2017 Guidelines Cohort

Of the 4,550 days, a similar breakdown of both days and patients below 0, <1%, <5%, <10%, and <15% PVCs is provided in Table 2.

**Table 2:** Days and Patients under certain PVC percentage burdens (2017 Guidelines Cohort). This table depicts how commonly there were days below certain PVC percentage thresholds, as well as how many patients had at least one day below certain PVC percentage thresholds.

| **% PVCs** | **0** | **<1%** | **<5%** | **<10%** | **<15%** |
| --- | --- | --- | --- | --- | --- |
| Days (of 4,550) | 1 | 214 | 538 | 1211 | 2244 |
| Days (%) | 0.02% | 4.81% | 12.09% | 27.21% | 50.43% |
| Patients (of 325) | 1 | 50 | 108 | 202 | 269 |
| Patients (%) | 0.31% | 15.38% | 33.23% | 62.15% | 82.77% |

### Days Out of 14 Surpassing ≥10,000/≥15% PVC Thresholds

#### 2014 Guidelines Cohort

The number of days out of 14 where the ≥10,000 PVC threshold is reached demonstrated a bimodal distribution (Figure 7a). Two significantly distinct subgroups were observed, accounting for 338/606 (55.8%) of patients – these groups were defined as “low frequency/low PVC” and “high frequency/high PVC”, referencing both how many days when the ≥10,000 PVC threshold was crossed, and how many PVCs were present on those threshold-crossing days. The “low frequency/low PVC” group included 214/606 (35.3%) of patients, with ≤3 of 14 days with ≥10,000 PVCs, with a mean of 12,342 ± 2,568 PVCs on those threshold-crossing days. The “high frequency/high PVC” group included 124/606 (20.5%) of patients, with all 14 days with ≥10,000 PVCs, with a mean of 24,580 ± 9,637 PVCs on those threshold-crossing days (p <0.001 comparing the groups).

**Figure 7a:**
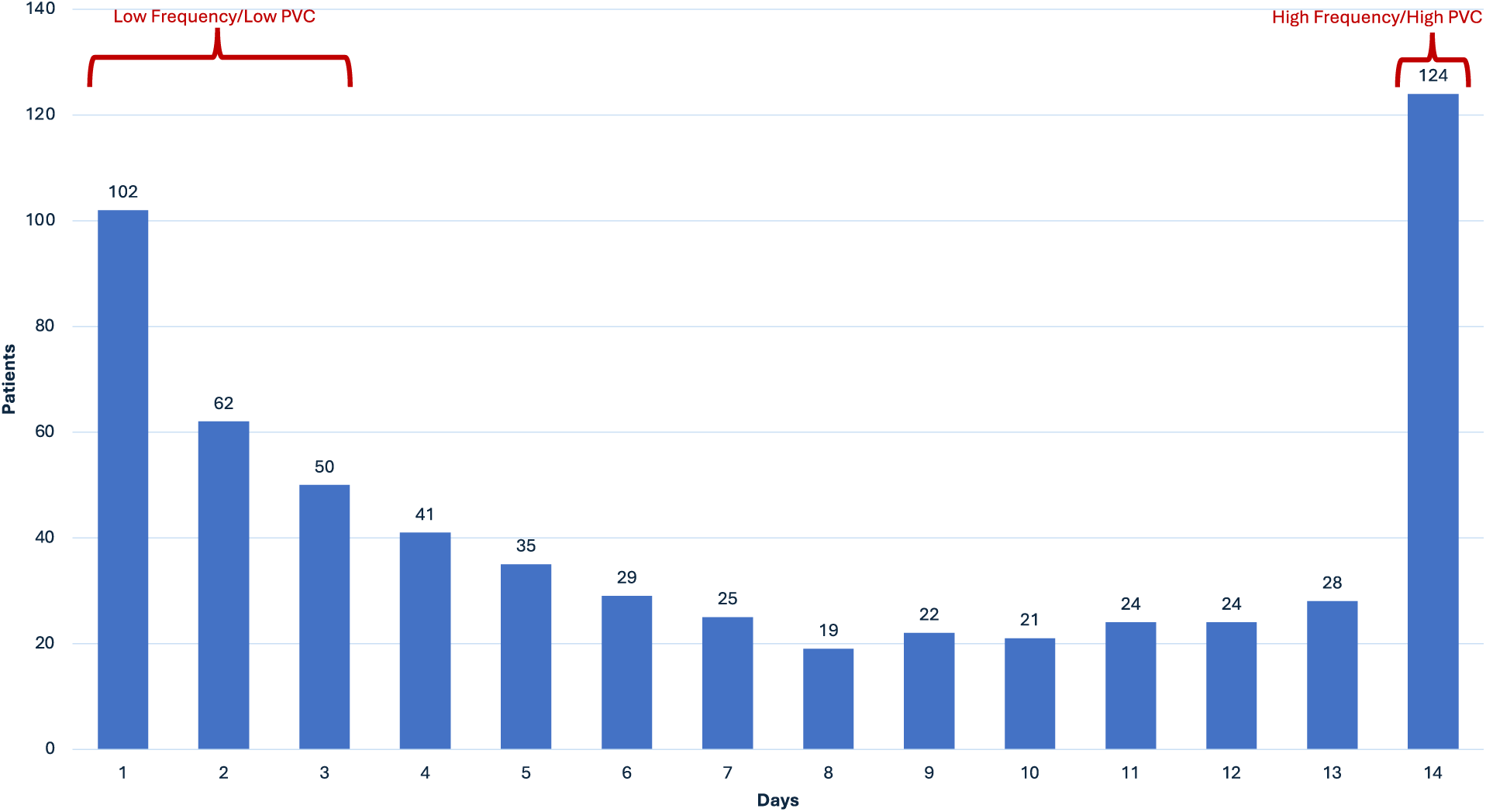
Total Number of Days Crossing 10,000 PVCs (2014 Guidelines Cohort) Bar Chart depicting the total number of days where ≥10,000 PVCs/day was reached (2014 Guidelines Cohort). A “low frequency/low PVC” and “high frequency/high PVC” bimodal distribution is demarcated, accounting for 338/606 (55.8%) of patients. “Low frequency/low PVC” accounts for 214/606 (35.3%) of patients with ≤3 of 14 days with ≥10,000 PVCs, with a mean of 12,342 ± 2,568 PVCs on those ≥10,000 PVC days. “High frequency/high PVC” accounts for 124/606 (20.5%) of patients with all 14 days with ≥10,000 PVCs, with a mean of 24,580 ± 9,637 PVCs on those days. The exact number of patients for each day number is noted above each bar.

#### 2017 Guidelines Cohort

Number of days out of 14 where the ≥15% PVC threshold is reached also demonstrated a bimodal distribution of significantly distinct subgroups including 169/325 (52.0%) of patients, with a “low frequency/low PVC” group (113/325 (34.8%) of patients, only ≤3 of 14 days with ≥15% PVCs, with a mean burden of 17.9 ± 2.8% PVCs on those days) and a “high frequency/high PVC” group (56/325 (17.2%) of patients, all 14 days with ≥15% PVCs, with a mean burden of 28.5 ± 7.0% PVCs on those days), p <0.001; Figure 7b.

**Figure 7b:**
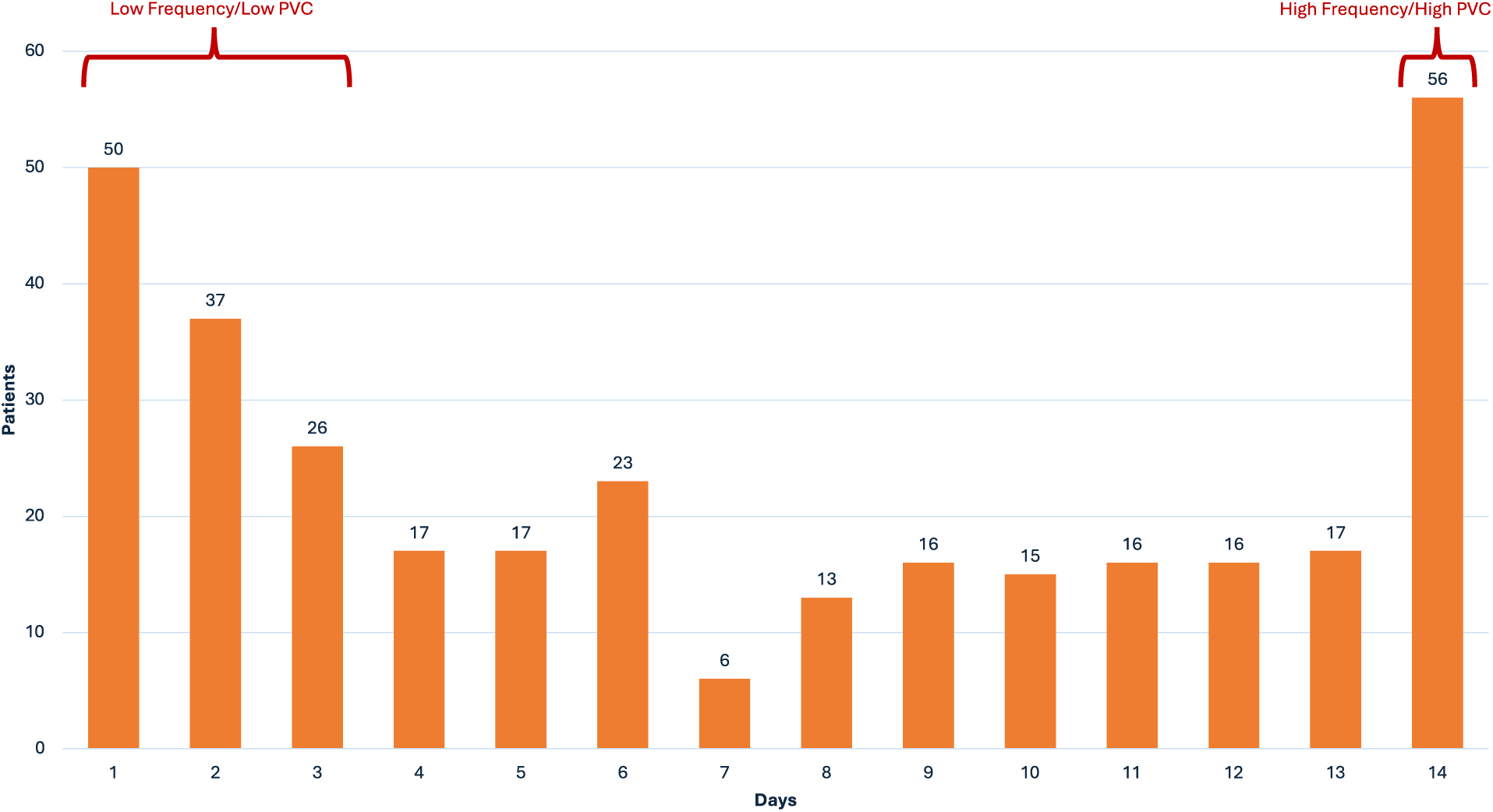
Total Number of Days Crossing 15% PVCs (2017 Guidelines Cohort) Bar Chart depicting the total number of days where ≥15% PVCs was reached (2017 Guidelines Cohort). A “low frequency/low PVC” and “high frequency/high PVC” bimodal distribution is demarcated, accounting for 169/325 (52.0%) of patients. “Low frequency/low PVC” accounts for 113/325 (34.8%) of patients with ≤3 of 14 days with ≥15% PVCs, with a mean burden of 17.9 ± 2.8% PVCs on those ≥15% PVC days (IQR 2.5%). “High frequency/high PVC” accounts for 56/325 (17.2%) of patients with all 14 days with ≥15% PVCs, with a mean burden of 28.5 ± 7.0% PVCs on those days. The exact number of patients for each day number is noted above each bar.

### Number of Days Needed to Reach ≥10,000/≥15% PVC Thresholds

#### 2014 Guidelines Cohort

317 patients (52.3%) met the ≥10,000 PVC/24h threshold within one day. A further 61 (10.1%) and 41 (6.8%) patients met the threshold in days 2 and 3, respectively. Following this, an additional 2-4% of new patients met the threshold with each additional monitoring day.

#### 2017 Guidelines Cohort

175 patients (53.9%) met the ≥15% PVCs/24h threshold within one day. A further 26 (8.0%), 24 (7.4%), and 25 (7.7%) patients met the threshold in days 2, 3, and 4 respectively. Following this, an additional 2-3% of new patients met the threshold with each additional monitoring day.

Figure 8 depicts these findings in both cohorts.

**Figure 8:**
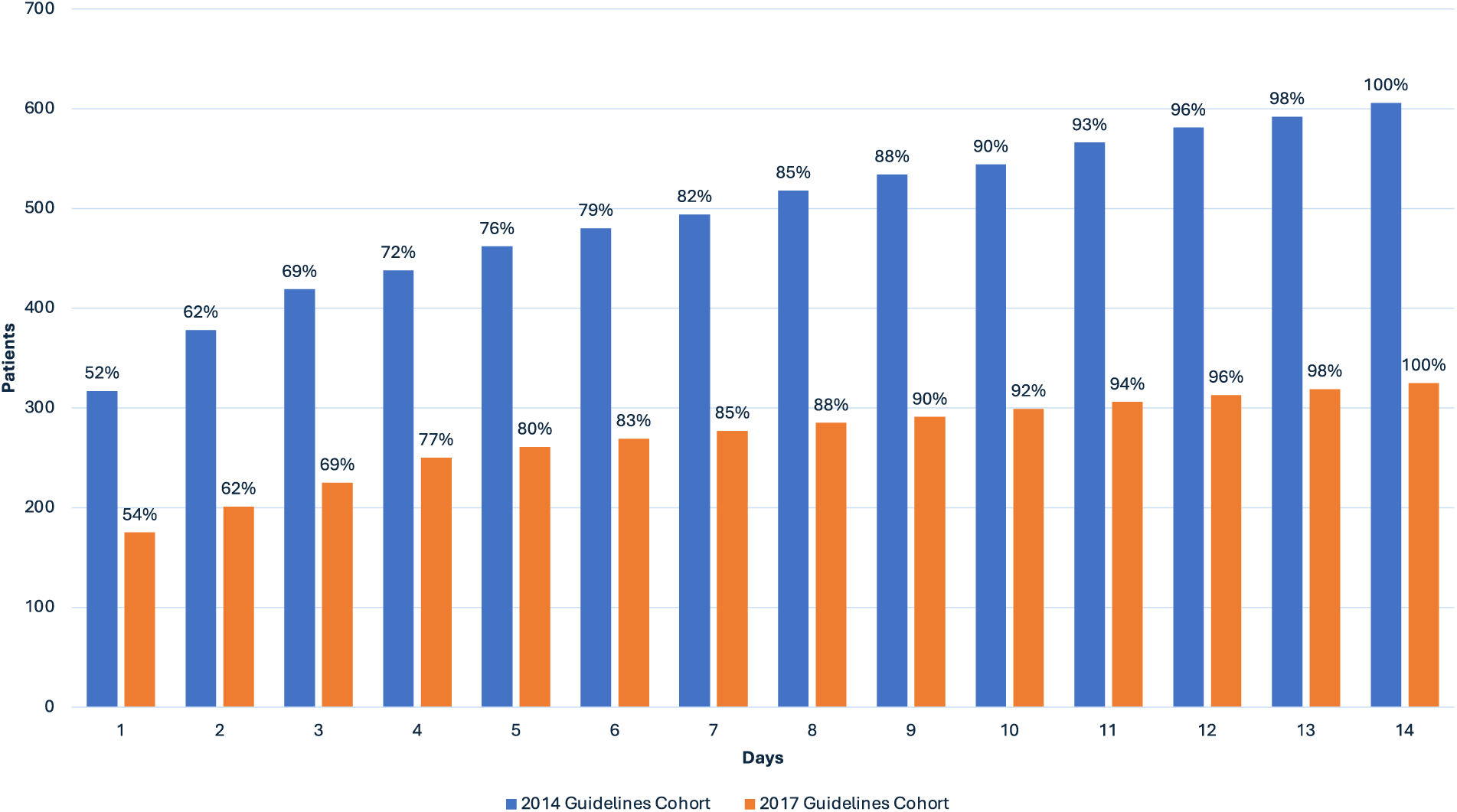
Consecutive Days Needed to Cross PVC Thresholds. Bar chart depicting the number of days needed to reach PVC thresholds. Blue bars represent the 2014 Guidelines Cohort (≥10,000 PVCs on at least one day), and orange bars represent the 2017 Guidelines Cohort (PVC burden ≥15% on at least one day). The percentage of patients within each respective cohort meeting the threshold on each day is shown above each bar.

### Influence of Tachycardia

The correlation between PVCs and total heart beats was assessed to gain insight into the influence of heart rate on PVC burden. Overall, daily PVC count and daily total heart beats were not well correlated (R^2^ = 0.045, Figure S1).

To look specifically at the relationship between tachycardia and PVCs, days were classified as “tachycardic” (mean heart rate ≥100 bpm) and “nontachycardic” (mean heart rate <100 bpm), with 131 “tachycardic” days and 8,353 “nontachycardic” days identified. The average daily PVC count of the “tachycardic” days (12,401 ± 11,349) and “nontachycardic” days (12,185 ± 9,415) was similar (p = 0.83). However, the average daily PVC percentage was higher on the “nontachycardic” days (11.6 ± 8.6%) compared to the “tachycardic” days (8.2 ± 7.6%, p <0.001).

Secondly, patients were equally classified into “tachycardic” and “nontachycardic”. 7 of the 606 patients were deemed “tachycardic” over the 14 day monitoring period with an average HR ≥100 bpm. The average daily PVCs of the “tachycardic” group (10,929 ± 2,142) was similar to the nontachycardic” group (12,202 ± 8,336; p = 0.21). However, the average PVC percentage of the “nontachycardic” group was higher than that of the “tachycardic” group (11.68 ± 7.55% vs. 7.32 ± 1.41%; p <0.001).

## DISCUSSION

The salient, novel findings of this large and highly detailed multi-day PVC burden analysis are that A) Daily PVC burden demonstrates significant inter- and intra-patient variability, even within a PVC-enriched cohort, B) The highest yield of patients passing guideline-suggested PVC thresholds is achieved by days 3-4, with longer-term PVC monitoring increasing sensitivity for detection, and C) Two previously non-identified PVC patterns meeting guideline suggested PVC-thresholds may exist (“high frequency/high PVC” and “low frequency/low PVC”). Similar findings were observed in both the 2014 Guidelines Cohort and the 2017 Guidelines Cohort.

### PVC Variability

Even amongst this PVC-enriched population, high intra-patient PVC variability is clearly demonstrated in this analysis. When looking at all patients, variable variability is observed - while the majority of maximum/minimum day fold change was between 2-10, >10k fold change was seen in 1.2% of patients over the 14-day monitoring period. The peak maximum/minimum fold change was 28,292 in a patient with a day of 0 PVCs and another day of 28,292 PVCs. Day to day fold change was also variably variable – over 85% of instances demonstrated <2 fold change, though the peak day to day fold change was 4,610, observed in a patient with 13,829 PVCs on one day, followed by 3 PVCs the next day. This highlights that a single day PVC assessment may be unable to accurately assess actual clinical PVC burden. PVC variability has been previously suggested by other studies.^10–13^ This detailed analysis provides definite quantification of the clinical PVC variability observed in a high PVC population.

### Monitoring Duration

This study also provides insight into more optimal durations for PVC monitoring. Just over 50% of patients met guideline-specified thresholds after 1 day (317/606, 52.3% in the 2014 Guidelines Cohort and 175/325, 53.8% in the 2017 Guidelines Cohort). An additional 16.8% (102/606) of patients in the 2014 Guidelines Cohort, and an additional 23.1% (75/325) of patients in the 2017 Guidelines Cohort met these thresholds by days 3 and 4, respectively. The sensitivity substantially drops off at this point to 2-4% of patients identified per day after this, but this 2-4% value persists without much diminishment through day 14.

This data can also be viewed in terms of days where guideline suggested cutoffs are not met. ∼50% of days in each cohort do not meet guideline cutoffs, equating to ∼80% of patients in each cohort having at least one of such days. Taken further, 5.8% of patient days in the 2014 Guidelines Cohort do not meet 1,000 PVCs and 4.8% of patient days in the 2017 Guidelines Cohort do not meet 1% PVCs, equating to 19.5% and 15.4% of patients, respectively, in each cohort having at least one of such days during monitoring. In this context, shorter duration monitoring may have not only missed expert consensus guideline-suggested PVC cutoffs, but even misclassified some patients as unlikely to suffer from a significant PVC burden altogether. Additionally, there were 3 patients in the 2014 Guidelines Cohort with at least one day of 0 PVCs and 1 patient in the 2017 Guidelines Cohort with at least one day of 0 PVCs, demonstrating that there is no “safe low number” of PVCs in a single 24 hr monitor which would predict that patients do not ever fulfill Guideline suggested criteria.

This is of particular interest as, in many studies looking at PVC suppression in those with left ventricular dysfunction, PVC thresholds were assessed by 24 hours of Holter monitoring. In their seminal paper, Duffee et al. saw improved clinical functional status and ejection fraction after pharmacologic PVC suppression in those with left ventricular dysfunction and >20,000 PVCs on a 24 hr Holter.^1^ Sarrazin et al. observed reversibility of left ventricular dysfunction with PVC ablation in those with ischemic cardiomyopathy and a >5% PVC burden (mean of 21.8 ± 12.5%) on 24 hr Holter.^3^ Ling et al. observed improvements in left ventricular dysfunction with medical and catheter-based PVC suppression in those with >6,000 RVOT PVCs (mean of 13,823 for antiarrhythmic group, 14,049 for catheter ablation group) on 24 hr Holter.^5^ It is certain that more patients meeting consensus guideline cutoffs will be identified through longer duration monitoring, though with increased sensitivity for detection the effect on overall outcome (response to PVC suppression/ablation) is difficult to predict and will require further investigation.

### PVC Subgroups

A novel finding in this study is the identification of two previously unrecognized subgroups of “low frequency/low PVC” and “high frequency/high PVC” within this PVC-enriched population, which correspond to the low and high bimodal peaks of days crossing the PVC thresholds during monitoring (Figure 7a, 7b). While both groups technically meet cutoffs for potential PVC suppression/ablation, it is physiologically plausible that the “high frequency/high PVC” subgroup may have a higher proclivity towards PVC-induced cardiomyopathy and/or a more robust response to PVC suppression/ablation (given that PVC cutoffs in this group were met not only more frequently, but also by a much higher margin). Differentiation of these two patient groups would require the extended monitoring performed in this study. Further study is required here to better characterize patient selection to determine who may benefit most from PVC suppression.

### PVCs and Tachycardia

The relationship between tachycardia and PVC burden is of additional interest. When looking strictly at the absolute number of PVCs, there is no significant difference between “tachycardic” and “nontachycardic” days or patients. When viewed through the lens of PVC percentage, “tachycardic” days/patients actually have a lesser PVC burden, implying that “tachycardic” days/patients are so due to more conducted beats and not more PVCs or complex VT. Animal models have demonstrated that RV apical paced-PVC burdens of 25% and 33% were capable of causing PVC-induced cardiomyopathy without the presence of tachycardia (HR >100 bpm) - in these models, a paced-PVC burden of 50% was required to produce tachycardia, a PVC burden which is not clinically observed.^14^ In fact, in our study there was not a single day of persistent bigeminy or persistent complex VT with PVC burden ≥50%. The overall peak PVC burden in a single day was 48.1%, and the overall peak PVC burden over the course of the 14 days of monitoring was 40.7% (both in the same patient). There were 69 total study days with a PVC burden ≥40%, and only 3 of these days had a HR ≥100 bpm. The aforementioned peak PVC burden patient had no days with a HR ≥100 bpm. Our findings thus support the notion that tachycardia plays a limited role in PVCs, and that tachycardia-mediated cardiomyopathy and PVC-induced cardiomyopathy are likely distinct etiologies.

#### Limitations

This study has several limitations. The patient demographics and reason for ambulatory rhythm monitoring were not provided – thus, the patient population here may not accurately reflect a real-world population of patients where rhythm monitoring is ordered for the explicit purpose of PVC analysis, particularly in the setting of suspected PVC-induced cardiomyopathy. PVC characteristics (such as morphology, QRS duration and coupling intervals) are also not known. The patient selection (at least one day of ≥10,000 PVCs) was chosen to reflect a commonly used clinical cut-off, though this may limit the generalizability of the findings. It is important to note that the primary selection criteria for the study population used the ≥10,000 PVC cutoff (2014 Guidelines Cohort), making this the primary analysis. The ≥15% PVC burden subcohort (2017 Guidelines Cohort) was derived from 2014 Guidelines Cohort, and thus represents a secondary analysis of this population. While the consistency of the findings between both cohorts suggests validity, this presents an important limitation when interpreting the 2017 Guidelines Cohort findings. A direct comparison between the two cohorts may not be appropriate.

## CONCLUSION

In this large and highly detailed analysis of 14 day heart rhythm monitoring data in a PVC-enriched population, several important findings were identified. PVC burdens were found to have high variability. The highest yield of patients passing guideline-suggested PVC thresholds is achieved by day 3-4, though there is some additional value in continuing monitoring to 14 days to achieve the maximum sensitivity. Novel “low frequency/low PVC” and “high frequency/high PVC” subgroups were identified, which may allow for better identification and treatment of patients with suspected PVC-induced cardiomyopathy. “Tachycardia” in this population generally did not appear to be caused by more PVCs, but instead by more conducted beats. These findings provide a better understanding of clinically observed PVC patterns and improve identification, treatment, and further study design for patients with frequent PVCs.

## Data Availability

All data referred to in the manuscript is de-identified and available for review.

## Non-standard Abbreviations and Acronyms

EHRA: European Heart Rhythm Association
HRS: Heart Rhythm Society
APHRS: Asia Pacific Heart Rhythm Society.
AHA: American Heart Association
ACC: American College of Cardiology
HRS: Heart Rhythm Society

## Acknowledgements

a) <u>Acknowledgements:</u> We acknowledge the aid of iRhythm in providing deidentified datasets of patients with a high PVC burden to analyze.

b) <u>Sources of Funding:</u> There are no sources of funding to disclose.

c) <u>Disclosures:</u> The data analyzed in this manuscript was acquired in a non-financial collaboration with iRhythm. Outside of this, there are no additional industry relationships among any authors to disclose.

## Supplemental Materials

**Figure S1:**
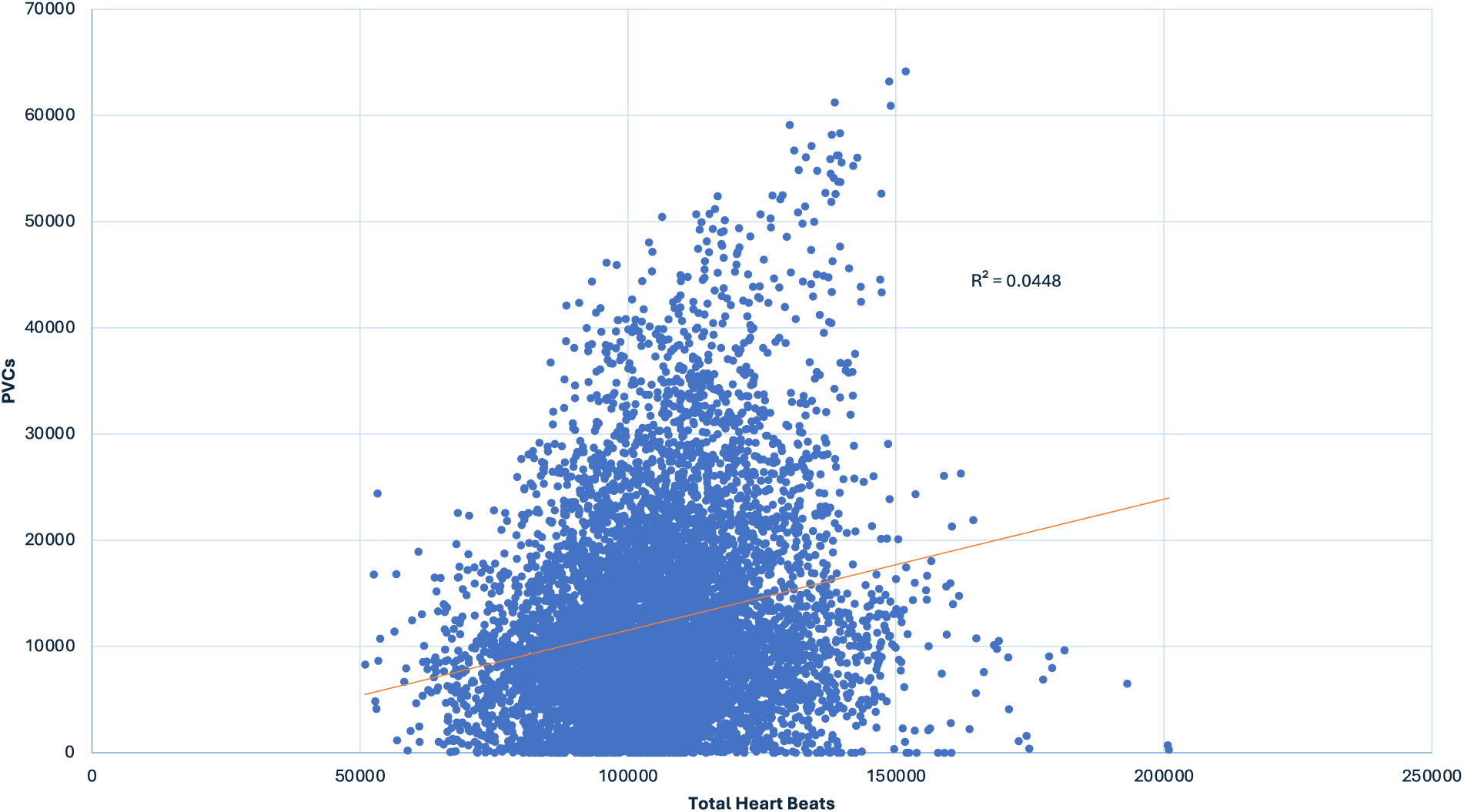
Correlation Between PVCs and Total Heart Beats. Scatterplot depicting the relationship between PVCs and total heart beats amongst all days analyzed (8,484 days). An R^2^ value of 0.045 indicates that PVCs and total heart beats are not well correlated.

## Notes

### Competing Interest Statement

The authors have declared no competing interest.

### Clinical Trial

This is a retrospective data analysis using de-identified patient data, not a prospective clinical trial.

### Author Declarations

Communication with the IRB of the University of Maryland School of Medicine confirmed that no special IRB approval was required given that only anonymous and deidentified data was used.

